# High ELF4 Expression in Human Cancers is Associated with Worse Disease Outcomes and Increased Resistance to Anticancer Agents

**DOI:** 10.1101/2020.05.16.20104380

**Authors:** Doris Kafita, Victor Daka, Panji Nkhoma, Mildred Zulu, Ephraim Zulu, Rabecca Tembo, Zifa Ngwira, Florence Mwaba, Musalula Sinkala, Sody Munsaka

## Abstract

The malignant phenotype of tumour cells is fuelled by changes in the expression of various transcription factors, which include some of the well-studied proteins such as p53 and Myc. Despite significant progress made, little is known about several other transcription factors, including ELF4, and how they help shape the oncogenic processes that occur in cancer cells. To this end, we performed a bioinformatics analysis to facilitate a detailed understanding of how the transcriptional variations of ELF4 in human cancers are related to disease outcomes and the cancer cell drug responses. Here, using mRNA transcription data of ELF4 from the Cancer Genome Atlas pan-cancer project of 9,350 samples, we identify two groups of patient’s tumours: those that expressed high ELF4 transcripts and those that expressed low ELF4 transcripts across 32 different human cancers. We uncover that patients segregated into these two groups are associated with different clinical outcomes. Further, we found that, besides being associated with comparatively worse disease outcomes, tumours that express high ELF4 mRNA transcript tend to be of a higher-grade, afflict a significantly older patient population and have a significantly higher mutation burden. By analysing dose-response profiles to 397 anti-cancer drugs of 612 well-characterised human cancer cell lines from the Genomics of Drug Sensitivity in Cancer, we discover that cell lines that express high ELF4 mRNA transcript are significantly less responsive to 129 anti-cancer drugs. Collectively our analyses have shown that, across the 32 different human cancers, the patients afflicted with tumours that overexpress ELF4 tended to have a more aggressive disease that is also is more likely more refractory to most anti-cancer drugs, a finding upon which we could devise novel categorisation of patient tumours, treatment and prognostic strategies.

## Introduction

Genetic alterations in several transcription factors lead to genome-wide transcription changes that drive the malignant phenotypes [1–3]. Although many of these transcription factors, such as ELF4 a member of E74-like factor family (ELF), play essential physiological roles in healthy proliferating cells, a series of genetic aberrations within genes coding for these factors result in oncogenic behaviour of healthy cells [4,5]. However, we know far less about how the transcription of ELF4 affects the disease aggressiveness across many human cancer types and their response to drug perturbations. Furthermore, despite the keen interest in determining how perturbations of ELF4 relate to specific aspects of malignant cells, compared to other transcription factors such as TP53 (PubMed score of 46333.87) and MYC (13294.24), ELF4 (35.41) remains among the least studied transcription factor in human cancers [6,7].

Analysis of ELF4 in various human cancers has revealed that it plays essential roles in cellular differentiation, proliferation, and apoptosis in cancer of the prostate and breast [8], and is paradoxically associated with oncogenic activity [3,8] and tumour suppressor roles [9]. Also, mutations in the coding sequence and transcriptional variations of ELF4 have been reported in various human cancers [3,5,10]. These and other recent studies [4,7,11–13] have additionally generated a new appreciation of how aberrant ELF4 influences the development of cancer, the anti-cancer drug response of tumours, and the disease treatment outcomes.

Our current understanding of ELF4 and other cancer genes has largely been facilitated by large cancer profiling projects such as The Cancer Genome Atlas (TCGA) [14] and the International Cancer Genome Consortium [15]. Data from these have guided the identification of frequently altered cancer genes and the cancer type-specific regulators [16–20]. Additionally, large-scale drug response screening projects, such as the Genomics of Drug Sensitivity in Cancer (GDSC) project [21], have been valuable in providing well-characterised cancer cell lines as models of disease for both drug discovery and the evaluation of drug action dependencies of cancer cells [22–24].

Here, we investigate the transcription variations of ELF4 across pan-cancer TCGA tumours to identify the changes that have meaningful clinical value. By linking this information with the drug response profiles from the GDSC, we also investigate variations in the response of cancer cell types that are associated with ELF4 transcription levels. Besides enabling the identification and selection of the most suitable anti-cancer drugs to treat a tumour with different ELF4 transcription signatures, a multiscale understanding of ELF4 across different cancer types will also likely yield better prediction of disease outcome.

## Results

### The expression of ELF4 transcript varies across human tumours

We obtained and analysed a TCGA pan-cancer dataset comprising the mRNA transcription levels of the ELF4 gene and comprehensively deidentified clinical information. The TCGA collected these datasets from 9,350 patients of 32 distinctive human cancers. We used the z-score normalisation methods to segregate the patient’s tumours into two groups of cancers: those cancers with higher levels of ELF4 (which we named as the "high-ELF4 tumours") and those with a lower level of ELF4 (“low-ELF4 tumours”) see Methods sections).

We examined whether the two groups of cancers were associated with different clinical outcomes. Remarkedly, using the log-rank test [25], we found that the duration of overall survival (OS) periods for the patients with high-ELF4 tumours (OS = 51.9118 months) were significantly shorter (p = 7.99e-64) relative to those of the patients with low-ELF4 tumours (OS = 114.673 months; Figure 1A). Correspondingly, we observed that the disease-free survival (DFS) periods were significantly shorter (log-rank test p = 1.07e-07) for the patients with high-ELF4 tumours than they were for the patients with low-ELF4 tumours (Figure 1B).

**Figure 1:**
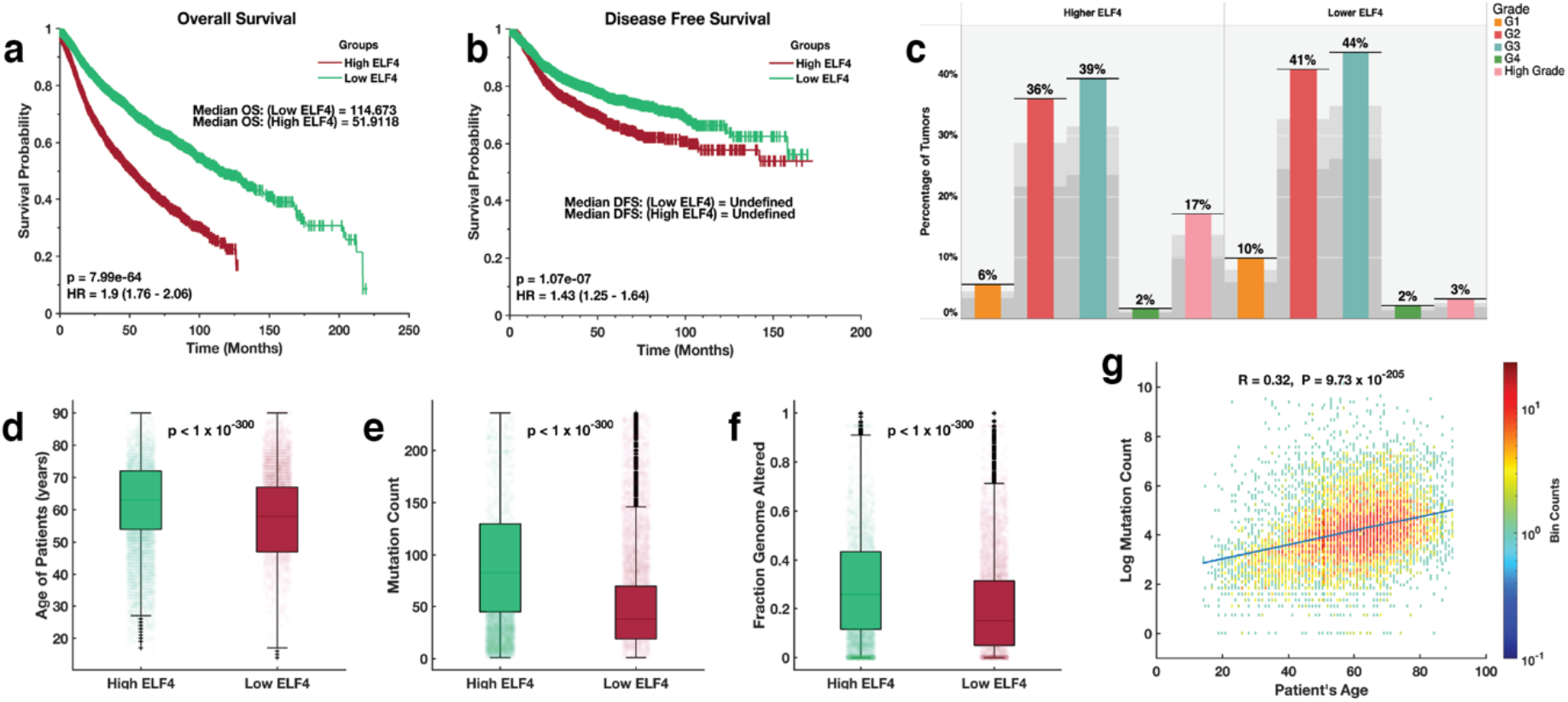
Kaplan-Meier curve of the overall survival periods **(a)** and disease-free survival periods **(b)** of TCGA patients with tumours that expressed high ELF4 and low ELF4 transcript levels. **(c)** the distribution of various grades tumours derived from 32 human cancer that expressed high ELF4 and low ELF4 transcript levels. Median comparison of the **(d)** age of the patients **(e)** number of mutations per patients’ tumours (f) the fraction of the genome altered for each patient tumour between the ELF4-high tumours and ELF4-low tumours. **(g)** binned scatter plot showing the Pearson’s linear correlation between the age of the patients and the number of mutations in each patient’s tumours. The data points spaced into rectangular bins and each point is coloured based on logarithm bin size with redder colours indicating a higher number of plots. The colour bar shows the colour scale.

Here, these findings expose an association between the transcription levels of ELF4 and the clinical outcomes across various cancer types.

### Disease factors associated with ELF4 expression

To understand why patients with tumours that express higher levels of ELF4 tended to have worse clinical outcomes (or a more aggressive disease). We assessed the distribution of various tumour grades across the two categories of tumours. Here, we found a significant association between the high-ELF4 tumours (Fisher exact test; odds ratio = 6.1, p = 4.5 x 10^-47^) with high-grade tumours (Figure 1c).

We found that the median age of the patients with tumours that expressed higher ELF4 was significantly higher (median = 63 years) compared to patients with tumours that expressed lower levels of ELF4 (median = 58 years), Wilcoxon rank-sum test (Z = 119.2, p < 1 x10^-300^; Figure 1d). We compared the median number of mutations per patient’s tumours across the two groups of disease. Here, we found a higher mutation burden in tumours that expressed higher levels of ELF4 (median = 83 mutations per tumour) than those that expressed lower levels of ELF4 (median = 38 mutations per tumour), Z = 119.2, p < 1 x10^-300^; Figure 1e. Correspondingly, we found a higher fraction of the genome altered in the high-ELF4 tumours (median = 0.2605 per tumour), than those of the low-ELF4 (median = 0.1499), Z = 200, p < 1 x10^-300^; Figure 1f.

To further explore the relationship between the age of patients and the mutation load of the tumours afflicting them, we measured the Pearson’s linear correlation coefficients between the age of the patients and the number of mutations found within their tumours. Here, we found that these were significantly positively correlated (Pearson’s correlation = 0.32; p = 9.27 x 10^-205^; Figure 1d).

Collectively these results show that high-ELF4 tumours, compared to the low-ELF4 tumours, tend to be of a higher-grade, afflict a significantly older patient population and have a significantly higher mutation burden.

### The effect of ELF4 transcription varies within cancer-type

Next, we compared the duration of the overall survival periods between patients with tumours that expressed high and low levels of ELF4 across the 32 cancers. Here, for nineteen cancer types, we found that the overall survival periods were shorter for patients with high-ELF4 tumours compared to the patients with low-ELF4 tumours (Figure 2a; Supplementary File 1). Among these nineteen cancer types are kidney renal clear cell carcinoma, kidney renal papillary cell carcinoma and liver hepatocellular carcinoma. Surprisingly, for six cancer types, we found that the overall survival periods were longer for patients with high-ELF4 tumours than those afflicted by low-ELF4 tumours. These included, among others, urothelial bladder carcinoma, and adrenocortical carcinoma. For another seven cancer types, including diffuse large b-cell lymphoma and kidney chromophobe, we could not establish the overall influence of the ELF4 expression, as the median OS periods were undefined (i.e., more than 50% of the patients were alive by the reporting time) for both the high-ELF4 cancer patients and the low-ELF4 cancer patients.

**Figure 2:**
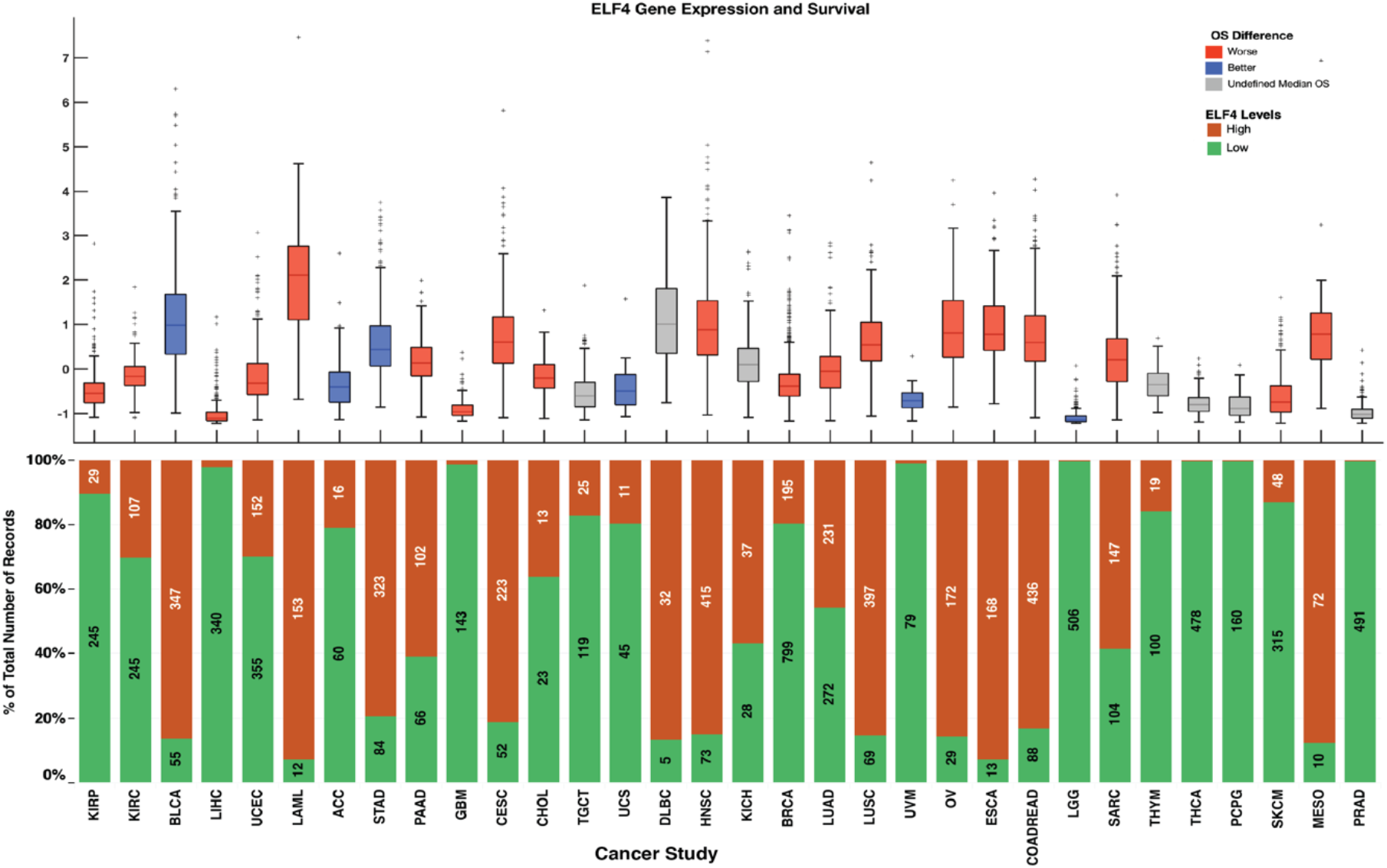
Top plot; overall survival comparison within cancer types between patient afflicted with high-ELF4 tumours and those afflicted with low-ELF4 tumours. Compared to the patient with low-ELF4 tumours, the red boxplots indicate median OS durations that are shorter for the patient with high-ELF4 tumours; the blue indicates median OS durations that are longer for the patient with high-ELF4 whereas the grey boxplots indicate median OS periods that are undefined (undefined median DFS period in that > 50% of patients survived beyond the study duration). On each box, the centre mark shows the median, and the left and right edges of the box show the 25th and 75th percentiles, respectively. The whiskers extend to the highest and lowest data points that are not outliers, and the outliers are shown individually using the ‘+’ symbol. The bottom plot shows the percentage of the total number of patients with tumours with each group given the colours of the bars. The number of patients with tumours labels the marks expressed high ELF4 and low ELF4 transcripts.

Furthermore, we explored the expression of ELF4 across tumour types across the two pan-cancer groups (high-ELF4 and low-ELF4). We found that ELF4 vary between cancer types, for example; brain lower-grade gliomas (99.9% of the tumours) and prostate adenocarcinomas (99.6%) tended to express lower ELF4 levels, whereas acute myeloid leukaemias (92.7%) and oesophageal adenocarcinomas (92.8%) tended to express higher levels of ELF4 (Figure 2b).

Overall, these findings show that while the expression of ELF4 is associated with increased disease aggressiveness of most cancer types, there are some exceptions.

### ELF4 expression is associated with significant resistance to most anti-cancer drugs

Since it is virtually impossible to test the primary patients’ tumours that either expressed high or low ELF4 levels to several anti-cancer drugs, we used cancer cell lines to test for any differences in their drug responses. From the GDSC database, we collected the dose-response of 612 cancer cell lines to 397 small molecule inhibitors [21], and their ELF4 mRNA transcription from the CCLE database [26]. We categorised the cell lines into two groups: 1) those that expressed high ELF4 mRNA transcripts (we refer to these as the “high-ELF4 cell lines”) and 2) those that expressed low ELF4 mRNA transcripts (the “low-ELF4 cell lines”; see Methods section). Here, the categories of the cancer cell lines that have ELF4 transcription levels that correspond to those of cancer cells from the patient groups could be used to examine how ELF4 transcript levels in the patient’s tumour cells are likely to influence the effectiveness of particular anti-cancer drugs.

We then compared drug z-score normalised half-maximal inhibitory concentration (IC50) values between the two categories of cells for 397 anti-cancer drugs.

Remarkedly, we uncovered differences between the high-ELF4 cell lines and low-ELF4 cell lines in their observed dose-response to 130 drugs (Figure 3a). Compared to the high-ELF4 cell lines, the low-ELF4 cell lines were significantly more sensitive to 127 anticancer drugs, including SB52334 (Welch test; p = 9.76 x 10^-08^), Nutlin-3a (p = 4.13 x 10^-07^), and CD532 (p = 7.53 x 10^-06^) (Figure 3b; Supplementary File 2). Astoundingly, the high-ELF4 cell lines were only significantly more sensitive than the low-ELF4 cell lines to three anti-cancer drugs (Figure 3b; Supplementary File 2). Furthermore, among the top thirty ranked significant anti-cancer drugs, the high-ELF4 cell lines were only significantly more sensitive (p = 1.16 x 10^-06^) than low-ELF4 cell lines to dasatinib; one anti-cancer drug (Figure 4).

**Figure 3:**
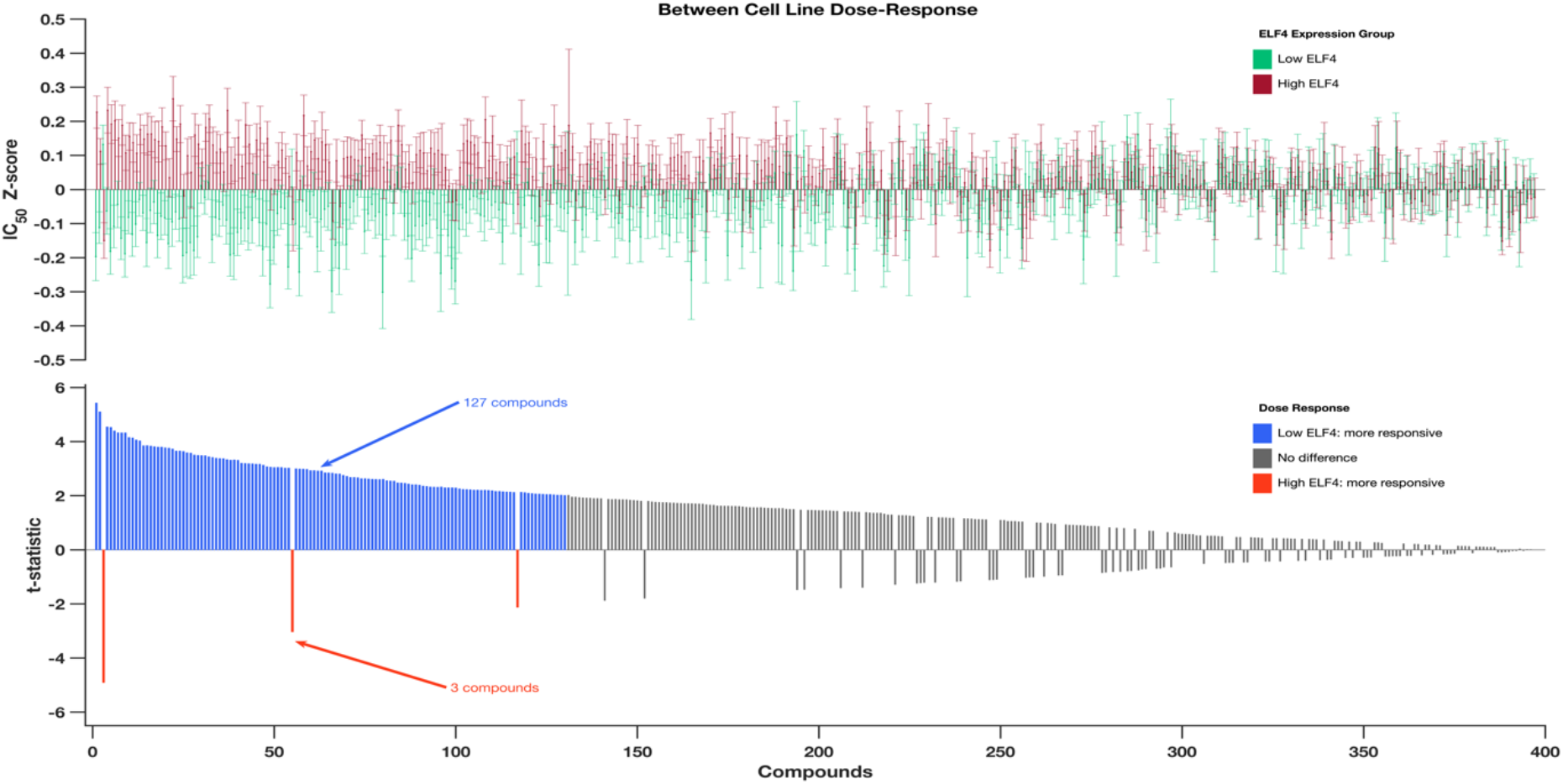
Top; Comparison of the dose-response profiles between the GDSC cancer cell lines that express higher ELF4 and those that express lower ELF4 transcript. The bar graphs show the logarithm IC50 values of the cancer cell lines that correspond to those that express higher ELF4 (red bars) and express lower ELF4 (green bar). Bottom; bar graph shows the t-value calculated using the Welch test for each of the 397 anti-cancer drugs used to profile the dose-response of the cell lines by the GDSC. The drugs along the columns are matched to the columns in the top plots. The orange bars indicate the drug to which the high-ELF4 cell lines are significantly more responsive; the blue bars indicate drug to which the low-ELF4 cell lines are significantly more responsive. The grey bars show drugs for which we found a non-statistically significant difference in the responsive between the high-ELF4 cell lines and low-ELF4 cell lines. For details refer to Supplementary file 2.

**Figure 4:**
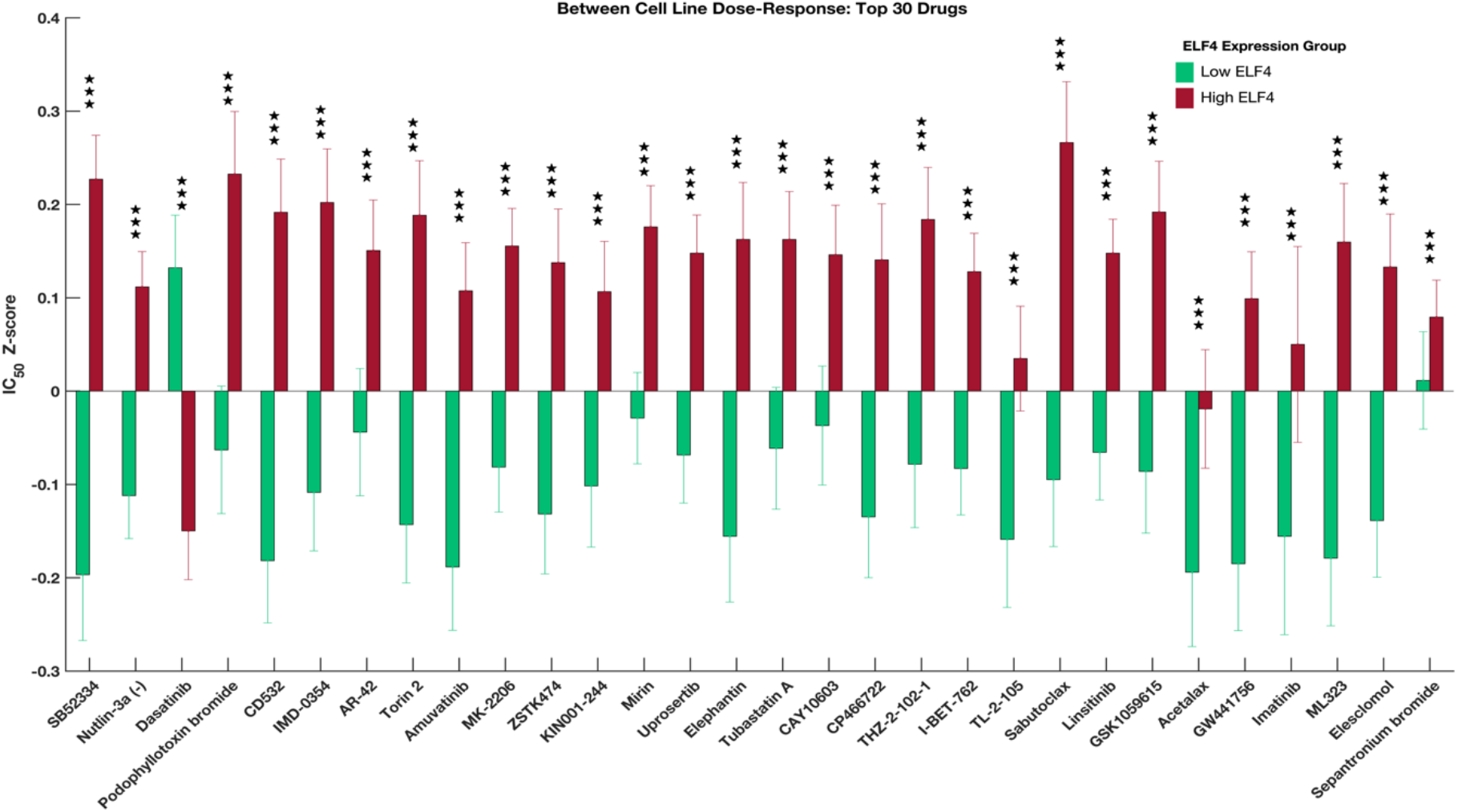
Top 30 drugs that showed the highest statistically significant dose-response differences between high-ELF4 cell lines and the low-ELF4 cell lines. The error bars show the standard error of the median logarithm IC50 value for each anti-cancer drug. The three stars show the degrees of statistical significance, i.e., p-values less than 0.001. For details, see Supplementary file 2.

Here, our results indicate that expression of ELF4 does not only detectably impact the clinical outcomes of the patient’s, but might also be a relevant variable to predict the response of tumours to various anti-cancer agents,

## Discussion

We examined the relationship between the transcription level ELF4 in thousands of primary tumours and cancer cell lines of 32 different cancer types and both the clinical outcomes and likely anti-cancer drug responses. Others have studied ELF4 in the context of tumorigenesis in cancers of the skin, breast and prostate [3,10,12,13]. To the best of our knowledge, our study is the first to characterise the consequences of ELF4 transcription variations across many distinct cancer types based on such many (9,350) primary tumours.

We showed that clinically relevant subtypes of patient tumours, across many cancer types, could be achieved by simply dividing the tumours into two categories based entirely on the transcription of ELF4: those tumours which express high ELF4 levels and those that express low ELF4 levels. Just as others have shown that the expression of various genes, including kinases such as the phosphoinositide 3-kinase and transcription factors such as p53, can have clinical implications [19,27–30], we show here that patients with low-ELF4 tumours tend to have significantly better clinical outcomes than patients with high-ELF4 tumours. Our results suggest that the tumour’s ELF4 transcription levels are directly correlated with the aggressiveness of cancer.

Our analyses indicate that high-ELF4 tumours are typical among the elderly, and these also tend to have a higher mutation load (Figure 1d, 1e and 1f). The frequency of somatic mutations increases with age in both non-cancerous and cancer tissue [31-34]; therefore we suggest that the link between the transcription of ELF4 and the mutation burden could be exploited to establish therapeutics that are more effective in treating specific cancers. Whereas ELF4 is virtually undruggable, we could employ a network analysis approach to identify the upstream regulatory proteins that could be targeted to treat tumours that overexpress ELF4 [35,36]. Furthermore, tumours with higher mutations are known to be more aggressive and respond significantly poorly to most anti-cancer drugs [37]. Accordingly, compared to the low-ELF4 tumours, we discovered higher gene mutation rates in high-ELF4 tumours, which could explain the poorer outcome exhibited by the patients afflicted with high-ELF4 tumours. Put together; we suggest that a combination of factors including the higher-grade tumours, higher mutation burden and the older patient population may in part explain why patients afflicted with high-ELF4 tumours level have worse survival outcomes.

There is a link between the expression of specific genes in cancer cells and the variations in the response of these cells to drug perturbation, and transcription levels of these genes can, thus, impact disease treatment outcomes [26,38–42]. With the advent of personalised medicine, growingly, the aim is to guesstimate the drugs to which a tumour is most likely responsive [43,44]. Here, to surmount the impractical barrier of testing hundreds of individual drugs on a specific tumour, cell lines that have genetic features resembling that of the tumour are convenient in inferring the drug responses of that tumour [21,26,45,46]. Suitably, by probing the drug response profiles of cancer cell lines, we showed that high-ELF4 tumours and low-ELF4 tumours perhaps respond differently to many anti-cancer drugs. More specifically, compared to the low-ELF4 tumours, the high-ELF4 tumours tended to be significantly responsive to only three out of 397 anti-cancer drugs. This finding implies that in aside the high-ELF4 tumours being significantly aggressive than low-EFL4 tumours, the high-ELF4 tumour patients may also exhibit worse clinical outcomes merely due to the refractory nature of the high-ELF4 tumours to most anticancer drugs. Additionally, since refractory tumours require more aggressive treatment protocols with higher drug doses, it would follow that patients with high-ELF4 tumours are likely to be exposed to such treatments, thus would tend to experience more adverse drug effects that might unfavourably impact their survival [47–50].

Altogether, we have shown that expression of ELF4 varies across tumours of the 32 cancer types investigated and that there is a link between the transcription levels of ELF4 in tumours to the age of patients, the mutation burden and the clinical outcomes. Further, our analyses of the dose-response profiles of well-characterised cell lines have revealed that the transcription levels of ELF4 in tumours may be related to the varied responses of the cancer cell to drug perturbation, a finding that could help shape precision medicine.

## Methods

We accessed and processed a pan-cancer TCGA project dataset derived from 9,350 patients afflicted with 32 distinct human cancers [51]. These datasets include evenly processed mRNA transcription data, gene mutation data, and comprehensive deidentified clinical data for all patients.

Then we segregated the patient’s tumours across the pan-cancer studies into two groups: those that expressed a higher level of ELF4 mRNA transcripts and lower level of ELF4 mRNA transcripts. To achieve this, first, we applied z-normalisation to the measured ELF4 mRNA transcript across all the tumours. Then we considered those tumours with ELF4 mRNA z-score > 0 to have high ELF4 levels (3887 tumours) and those tumours with ELF4 mRNA z-score < 0 to have low ELF4 levels (5463).

### Survival analysis

We used the clinical information provided by the Cancer Genome Atlas and the corresponding TCGA sample IDs to match the patient’s samples to the appropriate clinical outcomes and sample features. Then we used the Kaplan-Meier approach to estimate the duration of the overall survival and disease-free survival periods between tumours that expressed a high level of ELF4 mRNA transcripts and low levels of ELF4 mRNA transcripts [25]. Furthermore, across each of the 32 cancer types, we used the Kaplan-Meier method to compare the duration of the overall survival periods between patients afflicted with high-ELF4 tumours and those afflicted with low-ELF4 tumours (see Supplementary File 1).

### Cancer grades across tumours that expressed high and low levels ELF4

We matched the TCGA sample IDs to the TCGA IDs provided in the clinical sample information to match the patients with tumours that either expressed low or high ELF4 levels to the clinical features of the samples. Then we performed a Chi-squared test to compare the distribution of tumours of various grades across each category of tumours that either expressed high ELF4 level and low ELF4 level.

### Distribution of tumour expressed high ELF4 and low ELF4 across cancer types

Across each of the 32 TCGA pan-cancer analyses, we counted the absolute number of tumours that expressed high ELF4 transcript levels and those that expressed low ELF4 transcript levels. Then we also obtained the overall percentage of tumours that expressed higher or lower mRNA transcript levels of ELF4 across each of the cancer types.

### Comparison of median patients age, number of mutations and fraction of the genome altered across the tumour groups

Across the two groups of tumours that expressed high ELF4 and low ELF4, we obtained the age of each patient. Furthermore, we calculated the number of gene mutations in each tumour and the fraction of the genome altered in each tumour. Then we used the nonparametric Mann-Whitney U-test for equality of medians for groups to compare the median age, the median number of mutations per tumour and fraction of the genome altered per patient between each group [52].

### Dose-response of cancer cell lines that expressed high and low ELF4 levels

We obtained 244,656 dose-response profiles of 612 cancer cell lines to 397 anticancer drugs from the Genomics of Drug Sensitivity in Cancer (GDSC) database [21]. The cell lines within the GDSC database represent 31 different human cancers. Also, we obtained the mRNA transcription levels of the ELF4 genes in these 612 cancer cell lines from the Cancer Cell Line Encyclopaedia (CCLE; [26]). Using the mRNA transcription data from the CCLE, we used the approach described above (for the primary TCGA tumours) to segregate the cell lines into two groups: 1) those with high ELF4 transcript levels and 2) those with low ELF4 transcript levels. Then we used the Welch test to compare the mean differences in the z-score transformed IC50 values between the cell lines with high ELF4 levels and those with low ELF4 levels, for each class of the 397 anti-cancer drugs (also see supplementary file 2).

### Statistical analyses

All statistical analyses were performed in MATLAB 2020a. Fisher’s exact test was utilised to assess associations between categorical variables. The Welch test for normally distributed data and the Mann-Whitney U-test for non-normally distributed data were used to compare continuous variables. Here, the distributions of the data were assessed using the Kolmogorov-Smirnov test for goodness of fit [53]. Statistical tests were two-sided and considered significant at p < 0.05 with the Benjamini-Hochberg correction applied were multiple comparisons were involved.

## Data Availability

The data that support the findings of this study are available from the following repositories: cBioPortal (https://www.cbioportal.org/), Genomics of Drug Sensitivity in Cancer (https://www.cancerrxgene.org/), and the Cancer Cell Line Encyclopaedia (https://portals.broadinstitute.org/ccle/).

## Code Availability

All the MATLAB source code used to process and analyse is available from the corresponding author upon reasonable request.

## Ethics Approval

The study protocol was approved by the University of Cape Town; Health Sciences Research Ethics Committee IRB00001938. The analyses in this study utilised publicly available mRNA transcription and dose-response datasets and deidentified clinical information that were collected by the TCGA, CCLE and GDSC from consenting participants. Here, all analyses were performed following the relevant policies, regulations and guidelines provided by the TCGA, CCLE and GDSC for analysing their datasets and reporting of the findings.

## Author Contributions

The study was conceptualised by DK, PN, MZ, and VD; The formal methodology was devised by VD, DK, PN, MZ, EZ, RT, FM, ZN, SM, and MS; VD, DK, PN, FM, RT, ZN, EZ, SM, and MS performed the formal analysis of the datasets; VD, DK, MZ, RT, ZN, FM, EZ, SM, and PN wrote the draft manuscript; the manuscript was revised by DK, PN, MZ, RT, FM, VD, ZN, EZ, SM, and PN, and MS created visualisations.

## Description of Supplementary Files

Supplementary File 1: Results for within cancer types comparisons of the duration the overall survival for patients with tumours that expressed high ELF4 and expressed low ELF4 transcription levels.

Supplementary File 2: comparison of the dose-responses of the high-ELF4 and low-ELF4 cell lines for 397 anti-cancer drugs that were profiled by the GDSC.

